# Oxidative stress markers predict treatment outcomes in patients with generalized anxiety disorder treated with selective serotonin reuptake inhibitors

**DOI:** 10.1101/2024.09.07.24313247

**Authors:** Lijun Cui, Jingjing Lu, Zhongxia Shen, Jielin Zhu, Huanxin Chen, Shenliang Yang, Shikai Wang, Xinhua Shen

## Abstract

**Introduction:** The etiology of generalized anxiety disorder (GAD) has not been fully understood, and oxidative stress may potentially contribute to its pathogenesis. However, there is no published evidence concerning the possible influence of oxidative stress on antidepressant treatment outcomes. This study investigated the ability of oxidative stress markers to predict treatment outcomes in GAD patients treated with selective serotonin reuptake inhibitors (SSRIs).

**Methods:** One hundred-one GAD patients and 100 healthy controls (HCs) were included in this study. The 101 GAD patients were selected for treatment with escitalopram (n=52) or sertraline (n=49) for eight weeks. Hamilton Anxiety Rating Scale (HAM-A) assessments were conducted before and after treatment. The serum levels of eight oxidative stress makers, malondialdehyde (MDA), lipid hydroperoxides (LPO), superoxide dismutase (SOD), glutathione peroxidase (GSH-Px), catalase (CAT), cortisol, high-density lipoprotein (HDL), and nitric oxide (NO) were measured using enzyme-linked immunosorbent assays (ELISA) before and after SSRI treatment in GAD patients and at the time of HCs enrollment.

**Results:** The serum levels of MDA, cortisol, and LPO were higher in GAD patients than in HCs (all *p<.001*), while SOD, GSH-Px, and CAT were lower than in HCs (all *p<.001*). The baseline MDA, LPO, NO, and cortisol levels were positively correlated with anxiety severity, while GSH-Px was negatively correlated. After eight weeks of SSRI treatment, the GSH-Px levels increased, and MDA and LPO decreased (all *p<.05*). Alterations in MDA levels co-varied with changes in anxiety measures (all *p<.05*). The ability of the receiver operating characteristic (ROC) area of the baseline MDA levels to predict the SSRI endpoint treatment response was 0.804 (*p<.05*).

**Conclusion:** The pathogenesis of GAD might involve oxidative stress. Moreover, serum MDA levels might predict treatment response to SSRIs. However, more research is warranted to confirm these findings.

## 1. Introduction

Among anxiety disorders, GAD is a chronic mental disorder characterized by persistent, excessive anxiety. It can begin during childhood or adolescence, and lifetime prevalence is higher in high-income countries than in low- and middle-income countries (5% versus 1.5 to 3%)[1]. SSRIs represent the first-line psychopharmacologic treatments for GAD[2]. However, a high rate of treatment resistance is related to comorbidity, the lack of effective biomarkers for diagnosis, and evaluation of efficacy[3, 4].

Although several psychological, genetic, and biological factors have been suggested to explain the cause of GAD[5–7], its pathogenesis is poorly understood. Previous studies have demonstrated that oxidative stress, characterized by an excessive oxidation process or insufficient antioxidant capacity, results in a substantial accumulation of free radicals[8], which affects the normal function of neurons and even results in cell death (Figure 1), and is central in the pathophysiology of anxiety disorders[9, 10]. While the brain accounts for only 2% of the body’s mass, it utilizes 20% of the body’s total oxygen consumption, and its moderate antioxidant defenses make the brain more vulnerable to redox imbalances[11]. In patients with psychiatric disorders (such as depression and anxiety disorders) who experience chronic stress, the interaction between immunoinflammation and oxidative stress exacerbates disease progression[12]. These patients exhibit elevated mononuclear cell levels in their blood, including activated phagocytes, macrophages, T helper-1 cells, and 17-like cells that produce substantial amounts of cytokines and reactive oxygen species (ROS). The actions of ROS-RNS (reactive nitrogen species) on fatty acids and proteins cause the formation of new oxidation and nitro saturation-specific epitopes. This activates Toll-like receptor-mediated autoimmune responses (TLR2/TLR4), which exacerbates cellular dysfunction, affecting the central regulation of neurotransmitter release, causing decreased brain energy metabolism and neuroplasticity, and ultimately triggering a neurodegenerative process[13–15]. Various studies have demonstrated that the excessive ROS generated due to oxidative stress can induce damage to the mitochondrial DNA within neurons, ultimately leading to psychiatric disorders[16, 17].

**Figure 1.**
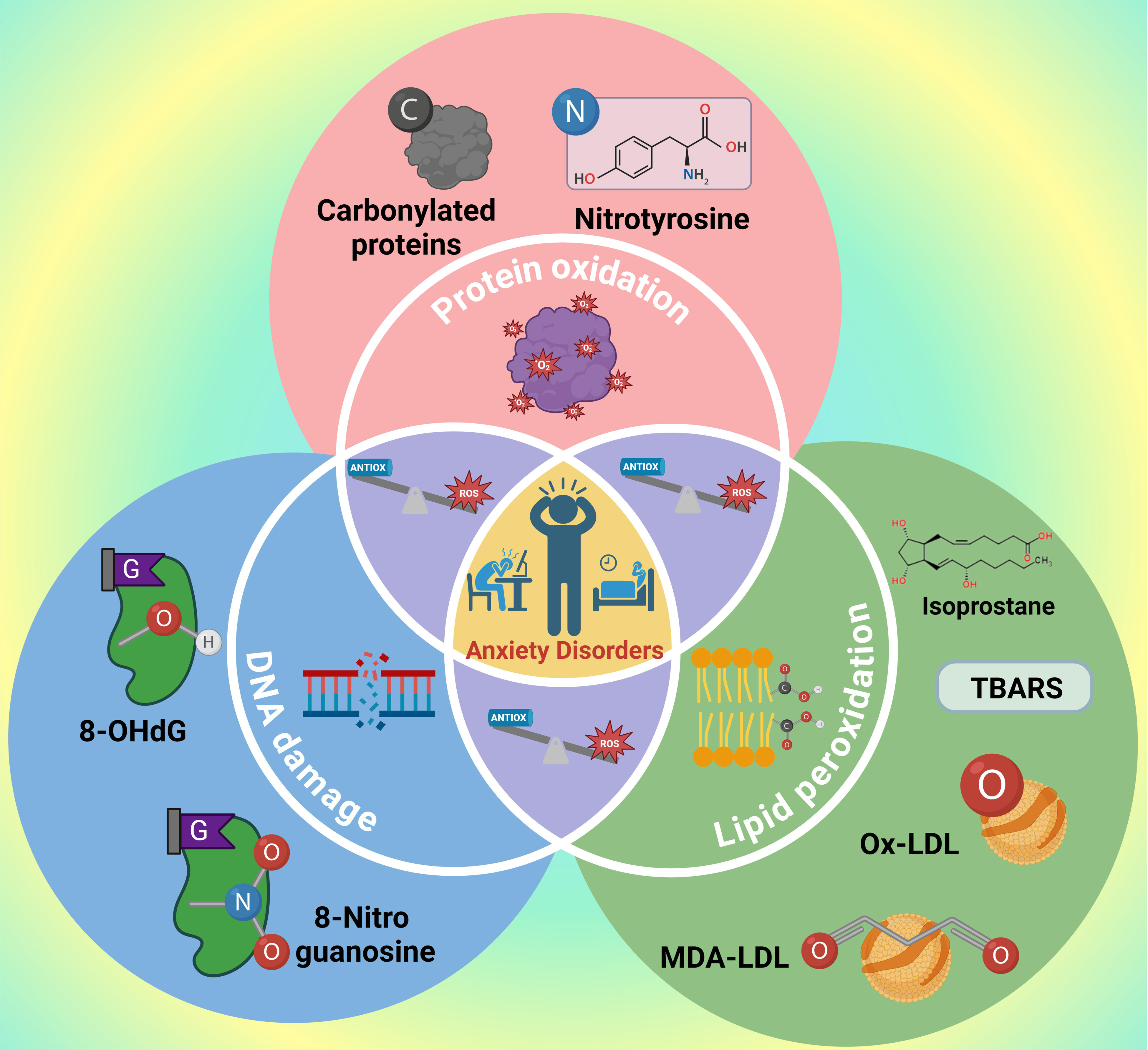
The impact of oxidative stress on cellular structural damage and its correlation with the initiation and progression of anxiety disorders.

Anxiety disorder is an adverse emotional state in which the body is constantly under stress, inducing an imbalance between oxidation and antioxidant systems[18]. The biomarkers LPO and MDA have been commonly employed to assess lipid peroxidation in animals and humans. MDA, the final product of lipid peroxidation, has the potential to induce cellular damage[19]. Psychological stress is associated with increased oxidative stress, and chronic stress can impair the normal feedback mechanism of the hypothalamic-pituitary-adrenal (HPA) axis, leading to chronically elevated cortisol levels, which are also oxidative stress markers[20]. Oxidative stress also induces alterations in the proteomics and lipidomics of HDL, leading to reduced HDL functionality, further impacting endothelial nitric oxide synthase (eNOS) activity. Consequently, there is a decline in NO synthesis, which plays a pivotal role in neuroprotection[21, 22].

The human body possesses several antioxidant defense mechanisms. SOD activity is the first line of defense against oxidative stress by catalyzing superoxide anion free radicals (O2^·−^) to H_2_O_2_[23]. The central redox molecule, H_2_O_2_, undergoes reduction to form H_2_O and O_2_ with the assistance of GSH-Px and CAT[24]. The impaired function of these antioxidants contributes to the initiation of anxiety disorders.

The quantification of oxidative stress levels commonly involves measuring antioxidant levels, antioxidant enzyme activities, and lipid peroxidation as a marker of oxidative damage. Findikli E. et al.[25] found significantly increased levels of MDA, and decreased levels of the antioxidant enzymes SOD and CAT in GAD patients.

These findings indicated that MDA and, to a lesser extent, SOD, might possess diagnostic significance, while CAT does not exhibit any diagnostic value.

Furthermore, researchers have suggested that regulation of cellular oxidative stress by cortisol correlates with neuroprotection; treatment with low-dose cortisol had neuroprotective effects, and treatment with high-dose cortisol was toxic to cortical neurons[26].

Thus, abundant evidence suggests that oxidative stress is involved in the pathogenesis of GAD[10, 27]. In particular, SSRIs have been shown to have anti-anxiety effects; previous reports indicated that sertraline increases neurogenesis and acts as an antioxidant[28]. Until now, there is no evidence that SSRI treatment alters the level of oxidative stress in GAD patients and whether oxidative stress biomarkers can predict treatment response to SSRIs. Therefore, this study examined eight oxidative stress markers associated with GAD: MDA, LPO, SOD, GSH-Px, CAT, HDL, NO, and cortisol. The primary aims of our study were: (1) compare the levels of these eight oxidative stress markers between GAD patients and HCs; (2) investigate potential correlations between the baseline levels of oxidative stress markers in GAD patients and HAM-A scores; (3) examine whether SSRI treatment affects the oxidative stress marker levels in GAD patients compared with before treatment; (4) determine whether oxidative stress marker changes were associated with improvements in anxiety symptoms; (5) identify whether baseline oxidative stress markers predicted the treatment response to SSRIs.

## 2. Materials and Methods

### 2.1 Participants

Recruitment of GAD patients was carried out by two psychiatrists at the Huzhou Third Municipal Hospital between November 2021 and February 2023, using structured diagnostic interviews utilizing the Diagnostic and Statistical Manual of Mental Disorders-5 (DSM-5) tool - SCID-5 to standardize the diagnostic process for mental disorders. All participants met the following criteria: (1) an age between 18-65, (2) a Hamilton Anxiety Rating Scale (HAM-A) score ≥17, and a 17-item Hamilton Depression Rating Scale (HAM-D17) score < 14, and (3) at the time of enrollment, all patients were free of any antidepressants and psychotherapy for at least two weeks. The following exclusion criteria were utilized: (1) the presence of other neurological or related medical disorders such as dementia, schizophrenia, bipolar disorder, and others; (2) subjects presented with immune-inflammatory disorders, including rheumatoid arthritis, chronic obstructive pulmonary disease, cancer, or type 1 diabetes; (3) patients were treated with antioxidant supplements such as vitamins and fish oil; (4) pregnancy. Concurrently, HCs were recruited from several communities; the HAM-A scores of the HCs all were ≤7. Any subjects with a history of chronic illness, psychiatric disorder, substance abuse, or antioxidant medication were excluded. Based on a pilot test, we calculated a necessary minimum sample size of 86 using PASS 15.0 with α=0.05 and β=0.10. The trial initially enrolled 110 GAD patients, and based on the 1:1 match ratio, 110 HCs were needed, but only 100 HCs were enrolled.

### 2.2 Drug treatment

A prospective cohort study design was used, in which GAD patients were treated orally with one of the SSRIs (escitalopram or sertraline) for eight weeks based on their clinical manifestations. The dose was adjusted according to the improvement of clinical symptoms and individual tolerance, and only one drug was taken during treatment. Finally, 55 participants took escitalopram, and 55 participants took sertraline. Other antipsychotics, psychotherapy, as well as drugs with potential antioxidant effects, such as anti-inflammatories, statins, neuroprotectants, and steroid hormones, were not allowed during the entire course of this study. The evaluators were blinded to the medication given to each patient. The use of benzodiazepines was prohibited, and hypnotics such as zolpidem were used to treat insomnia if needed, but for no more than two weeks.

### 2.3 Oxidative stress marker measurement

Blood was collected from GAD patients at the baseline (before treatment) and eight weeks after treatment. HCs blood was collected upon recruitment. Specifically, 10 ml of fasting venous blood was obtained from the cubital vein either on the day of recruitment or between 7 am and 9 am the following day to mitigate potential confounding effects from biological rhythm fluctuations of the measured factors.

Blood samples were centrifuged at 3,000 rpm for 10 min, and 5 ml of serum was placed in a −80 [refrigerator for further analysis. The serum levels of MDA, LPO, SOD, GSH-Px, CAT, cortisol, HDL, and NO levels were measured using ELISA. The ELISA kits were provided by the Jiangsu Jingmei Biotechnology Co., LTD. The serum samples were subjected to duplicate testing to calculate the mean value. The catalog numbers of the kits were as follows: (MDA:20230308X1, LPO:20230308N1, SOD:20230308R1, GSH-Px:20230308T2, CAT:20230308V1, cortisol:20230308E2, HDL:20230308Q1, and NO:20230308L1). Standard curves were established using standards for each compound, following the manufacturer’s instructions. The coefficient of variation for inter-test and intra-test comparisons of all test items was less than 10%.

### 2.4 Questionnaire used to measure anxiety

The severity and range of GAD symptoms in all patients were rated using the HAM-A by a single-blinded assessor throughout the study. The HAM-A was assessed at the initial baseline time point and eight weeks after treatment. The HAM-D 17 was used to evaluate depression at the same time points. The enrollment criteria for GAD patients included a depressive symptom score<14, aimed to manage comorbid depression in this population effectively. The treatment response was defined as a ≥50% reduction in the HAM-A score after eight weeks of treatment compared with the baseline HAM-A score. Patients were considered to be in remission if the HAM-A scores were≤7[29].

### 2.5 Statistical Analysis

Statistical analysis was performed using SPSS 19.0. The Shapiro-Wilk test was employed to assess the normality assumption of the data. The normal distribution of age, BMI, years of education, age of onset, HAM-A scores, HAM-D scores, and serum oxidative stress markers levels were expressed as means ± SD. The disease course of GAD patients was not normally distributed and was represented by the median (P25, P75), while the categorical variables of sex and family history of psychiatric disorders were depicted as frequency (N) and percentage (%). Paired sample t-tests were used to compare each oxidative stress marker’s pre- and post-treatment levels. A two-way ANOVA was employed to examine alterations in oxidative stress markers pre- and post-treatment with two SSRIs. The Pearson test was used to examine the correlation between HAMA-A scores and levels of oxidative stress markers in GAD patients at baseline, as well as the changes in HAM-A scores and oxidative stress marker levels after eight weeks of treatment with SSRIs. Logistic regression analysis and receiver operator characteristics (ROC) curves were applied to explore the predictive value of baseline oxidative stress marker levels in the treatment response. The *p*-values obtained from the *t*-tests, two-way ANOVA, and correlation analysis were corrected for FDR (false discovery rate) to make the statistical results more accurate and control for the potential increase in Type I errors due to multiple comparisons. Significance was set at *p*<.05 with adjustments for FDR.

## 3. Results

### 3.1 Baseline demographic and clinical characteristics of GAD and HCs

After eight weeks of treatment, five GAD patients (escitalopram=two; Sertraline=three) dropped out due to adverse events, two patients treated with sertraline were discharged early, and two patients (escitalopram=one; sertraline=one) withdrew due to poor treatment compliance. Thus, 101 patients (escitalopram=52; sertraline=49) and 100 HCs completed the study. There were no significant differences in sex, age, years of education, and BMI between GAD patients and HCs. However, the proportion of GAD patients with a family history of psychiatric disorders was significantly higher than in the HCs. The serum MDA, cortisol, and LPO levels in GAD patients were higher compared to HCs, and SOD, GSH-px, and CAT levels were lower than HCs. However, the HDL and NO levels were not significantly different between the two groups (all *p>.05*, see Table 1).

**Table 1.** Baseline demographic and clinical characteristics of GAD and HC.

| characteristics | GAD<br>(101) | HCS<br>(100) | $\chi^2/t$ | p-value |
| --- | --- | --- | --- | --- |
| Sex (male/female) | 46/55 | 41/59 | 0.423 | 0.516 |
| Age (years) | 48.43±12.00 | 45.72±8.67 | 1.834 | 0.068 |
| BMI (kg/m<sup>2</sup>) | 22.71±3.15 | 21.54±3.21 | 1.303 | 0.193 |
| Education (years) | 10.84±3.67 | 10.65±3.81 | 0.314 | 0.754 |
| Family history of<br>psychiatric disorders<br>(yes/no) | 14/87 | 4/96 | 5.993 | <0.05* |
| Age of onset (years) | 38.35±9.67 | NA | NA | NA |
| Duration of illness (months) | 84 (48,144) | NA | NA | NA |
| Baseline HAM-A score | 22.02±3.51 | NA | NA | NA |
| Baseline HAM-D 17 score | 8.90±2.12 | NA | NA | NA |
| MDA ( nmol/ml ) | 15.51±3.61 | 6.49±1.11 | 23.03 | <0.001*** |
| SOD ( pg/mL ) | 193.09±43.94 | 249.02±41.75 | -7.649 | <0.001*** |
| HDL ( mg/dL ) | 62.75±14.21 | 63.56±15.37 | -0.319 | 0.750 |
| GSH-Px ( pg/mL ) | 237.54±59.59 | 355.86±87.31 | -8.694 | <0.001*** |
| CAT ( pg/mL ) | 745.81±187.45 | 875.11±135.49 | -4.851 | <0.001*** |
| Cortisol ( pg/mL ) | 1373.72±342.23 | 999.27±199.05 | 8.631 | <0.001*** |
| NO ( μmol/ L ) | 230.41±75.51 | 223.94±54.51 | 0.694 | 0.571 |
| LPO ( nmol/mL ) | 121.68±25.27 | 101.52±19.43 | 3.816 | <0.001*** |

**Note.** BMI, Body Mass Index
The *p*-values obtained from comparison of oxidative stress marker levels at baseline between GAD patients and HCs were corrected by FDR. \**p*<.05, \*\*\**p*<.001

### 3.2 Correlation analysis between anxiety symptoms and oxidative stress markers at baseline

The baseline HAM-A score was positively correlated with the baseline levels of MDA, LPO, and cortisol. The baseline HAM-A score was negatively correlated with the baseline level of GSH-Px. The CAT, HDL, NO, and SOD levels were not significantly associated with anxiety symptoms (all *p>.05*, see Table 2).

**Table 2.** Correlation of the HAM-A scores and level of oxidative stress markers at baseline.

| Variables | HAM-A scores |  |
| --- | --- | --- |
|  | r | p |
| MDA(nmol/mL) | 0.349 | =0.024<sup>*</sup> |
| LPO(nmol/mL) | 0.470 | <0.001<sup>***</sup> |
| GSH-Px(pg/mL) | -0.458 | <0.001<sup>***</sup> |
| CAT(pg/mL) | -0.256 | =0.114 |
| HDL ( mg/dL ) | -0.142 | =0.177 |
| NO ( μmol/ L ) | -0.150 | =0.177 |
| SOD(pg/mL) | -0.097 | =0.498 |
| Cortisol(pg/mL) | 0.367 | =0.021<sup>*</sup> |

**Note.** *p*-values obtained by Pearson correlation analysis between HAM-A score and oxidative stress marker levels at baseline in GAD patients were corrected by FDR, *\*p*<.05, *\*\*p*<.01, *\*\*\*p*<.001

### 3.3 Comparison of oxidative stress markers and the HAM-A scores pre-and post-SSRI treatment

Six different oxidative stress markers between GAD patients and HCs were selected for additional analysis in GAD patients before and after eight weeks of treatment with SSRIs. The results revealed significantly decreased levels of MDA and LPO, while GSH-Px was significantly increased compared with the baseline (all *p<.001*, see Table 3). There were no significant differences in CAT, SOD, and cortisol levels pre- and post-treatment (all *p>.05)*. Meanwhile, the HAM-A score was statistically lower than the baseline after eight weeks of treatment (*p<.001*).

**Table 3.**
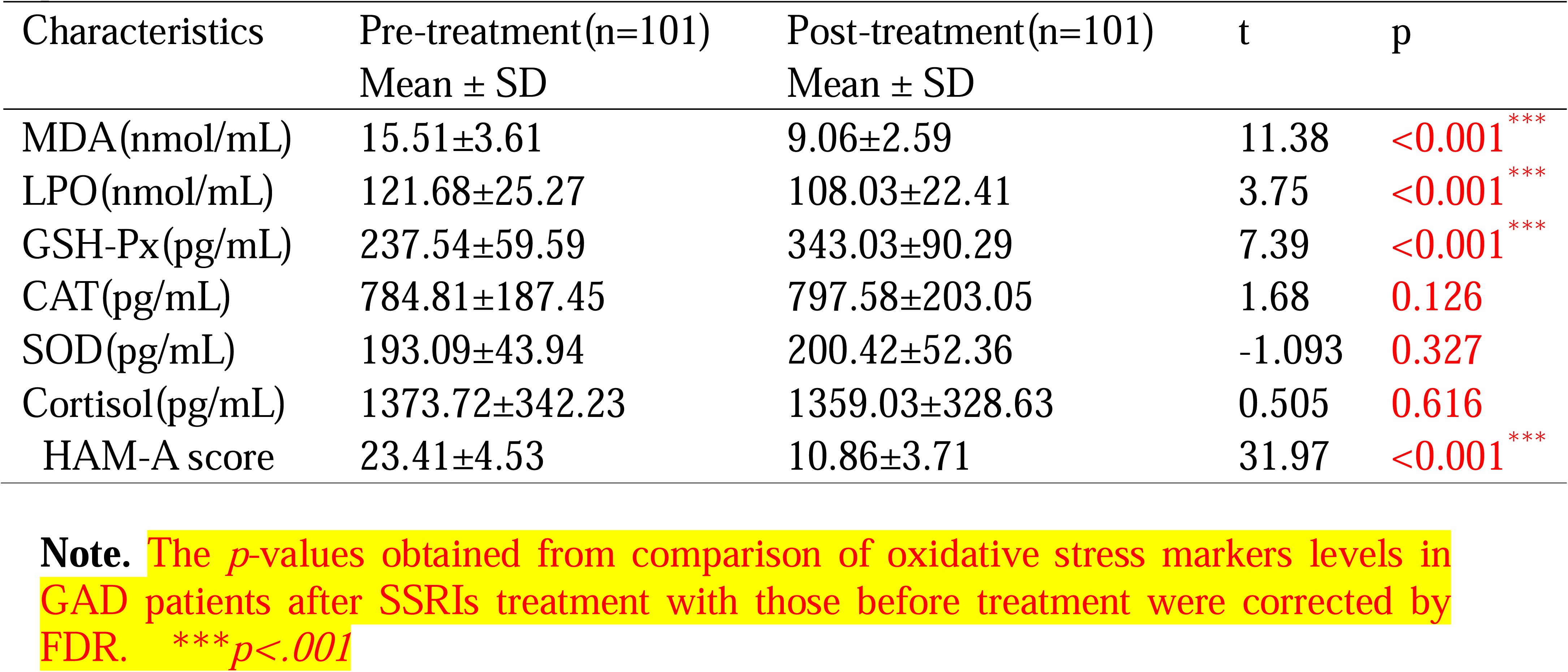
Measures of oxidative stress markers and the HAM-A scores pre-and post-treatment of SSRIs.

After conducting a subgroup analysis of the SSRIs, eight weeks of treatment with escitalopram and sertraline, no statistically significant differences were observed in the eight-week treatment remission rate (escitalopram 53.8% vs. sertraline 53.1%), and treatment response rate (escitalopram 78.8% vs. sertraline 81.6%) (all *p>.05*). The levels of serum MDA and LPO in GAD patients were decreased compared to pre-treatment levels (*p*<*.001, p*<*.01;* see Figure 2), while the levels of GSH-Px were elevated (*p*<*.001*). However, no significant difference was detected in the levels of oxidative stress markers between the groups receiving escitalopram and sertraline (all *p>.05*).

**Figure 2.**
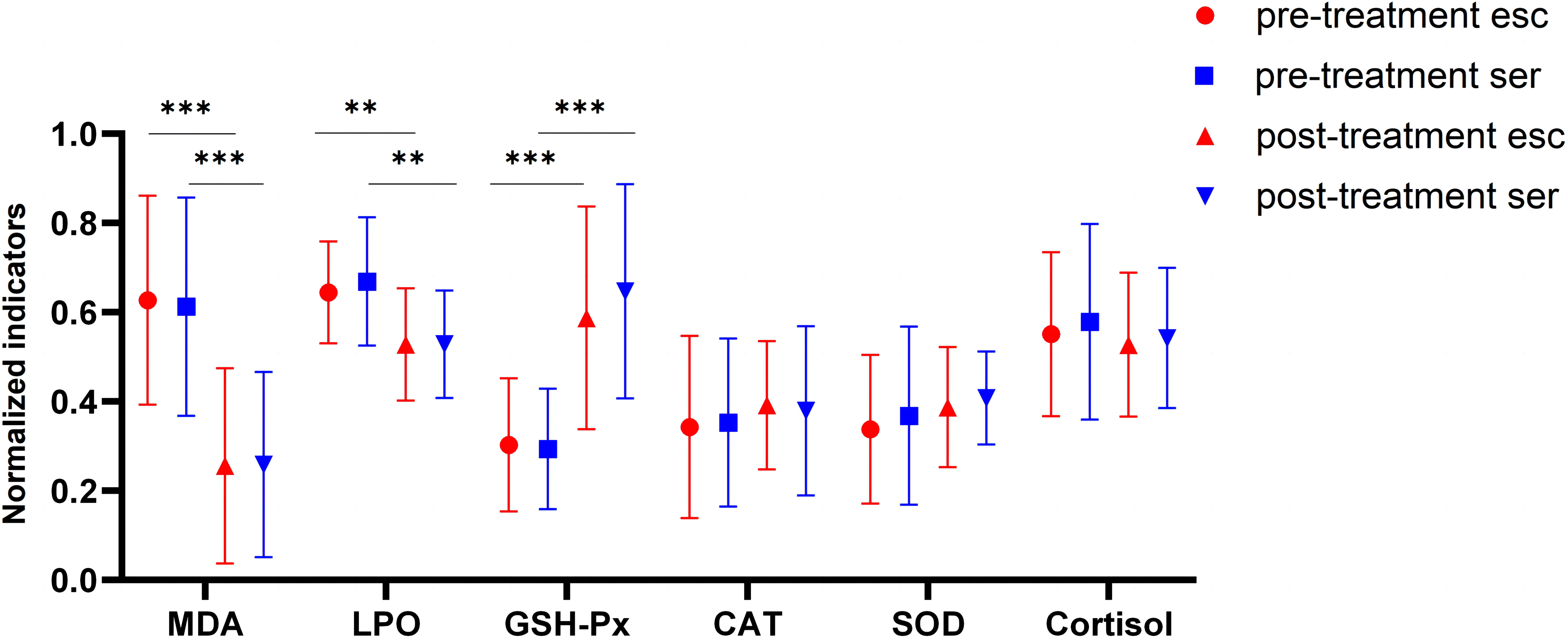
Comparison of oxidative stress markers in GAD patients pre- and post-treatment with escitalopram and sertraline. The levels of oxidative stress markers were normalized with a rescaling function. The *p*-values obtained from two-way ANOVA comparing pre-and post-treatment were corrected using FDR, \*\**p*<.01, \*\*\**p*<.001.

### 3.4 Correlation analysis between changes in anxiety symptoms and levels of oxidative stress markers

After eight weeks of treatment, the changes in MDA were positively correlated with reductions in HAM-A scores (*p<.05*; see Table 4). However, there was no correlation between changes in the GSH-Px or LPO levels and the reduction in HAM-A scores (*p>.05*). These results suggested that patients who experienced greater reductions in anxiety also exhibited larger decreases in MDA levels.

**Table 4.** Correlation analysis of the reduction in HAM-A scores and changes in oxidative stress markers pre-and post-SSRIs treatment.

| Characteristics | □ HAM-A |  |
| --- | --- | --- |
|  | r | p |
| □ MDA | 0.331 | 0.034* |
| □ LPO | 0.279 | 0.061 |
| □ GSH-Px | -0.195 | 0.171 |

**Note.** □: Pre- and post-treatment of SSRIs; the reduction in HAMA scores and changes in oxidative stress markers.
*p*-values obtained by pearson analysis of correlations between HAM-A reduction and oxidative stress marker levels in GAD patients after eight weeks of SSRI treatment were corrected by FDR, \* $p < .05$

### 3.5 The roles of baseline oxidative stress markers in predicting treatment response to SSRIs

Treatment response was defined as a reduction of HAM-A score ≥ 50% after SSRI treatment. However, a lack of treatment response was defined as a HAM-A score < 50%. Ultimately, 81 participants achieved a treatment response, and 20 did not. Regression models revealed that after controlling for sex, BMI, age, years of education, and duration of illness, the baseline concentration levels of MDA (*OR*=0.447, *95% CI*=0.332-0.968, *p=.034*) exhibited a significant predictive value for treatment response (see Table 5). Patients with lower baseline MDA levels responded better to treatment.

**Table 5.** Logistic regression analysis of baseline oxidative stress markers for treatment response to SSRIs.

| Variables | $\beta$ | S.E. | Wald | P | OR | 95% (CI) | |
| --- | --- | --- | --- | --- | --- | --- | --- |
|  |  |  |  |  |  | Lower | Upper |
| Sex | 1.272 | 0.809 | 1.718 | 0.132 | 3.304 | 0.551 | 8.805 |
| BMI (kg/m<sup>2</sup>) | -0.163 | 0.161 | 0.873 | 0.359 | 0.861 | 0.631 | 1.178 |
| Age (years) | -0.061 | 0.043 | 0.493 | 0.464 | 0.675 | 0.873 | 1.245 |
| Education (years) | 0.144 | 0.135 | 1.138 | 0.286 | 1.155 | 0.886 | 1.505 |
| Duration of illness (months) | -0.019 | 0.007 | 3.349 | 0.045* | 0.583 | 0.948 | 1.005 |
| MDA (nmol/mL) | -0.436 | 0.206 | 4.479 | 0.034* | 0.447 | 0.332 | 0.968 |

**Note.** Treatment response is the dependent variable, labelled as ‘0’ for no treatment response and ‘1’ for treatment responsive, and MDA is independent variables. Reference categories: sex, male (controlling for sex), BMI, age, years of education, and duration of illness. \*indicates $p < .05$ .

The predictive power of MDA was analyzed by receiver operating characteristic (ROC) curves. The ROC area of the baseline MDA levels for predicting endpoint treatment response was 0.885, indicating that MDA could be a significant predictor of treatment response to SSRIs (see Figure 3).

**Figure 3.**
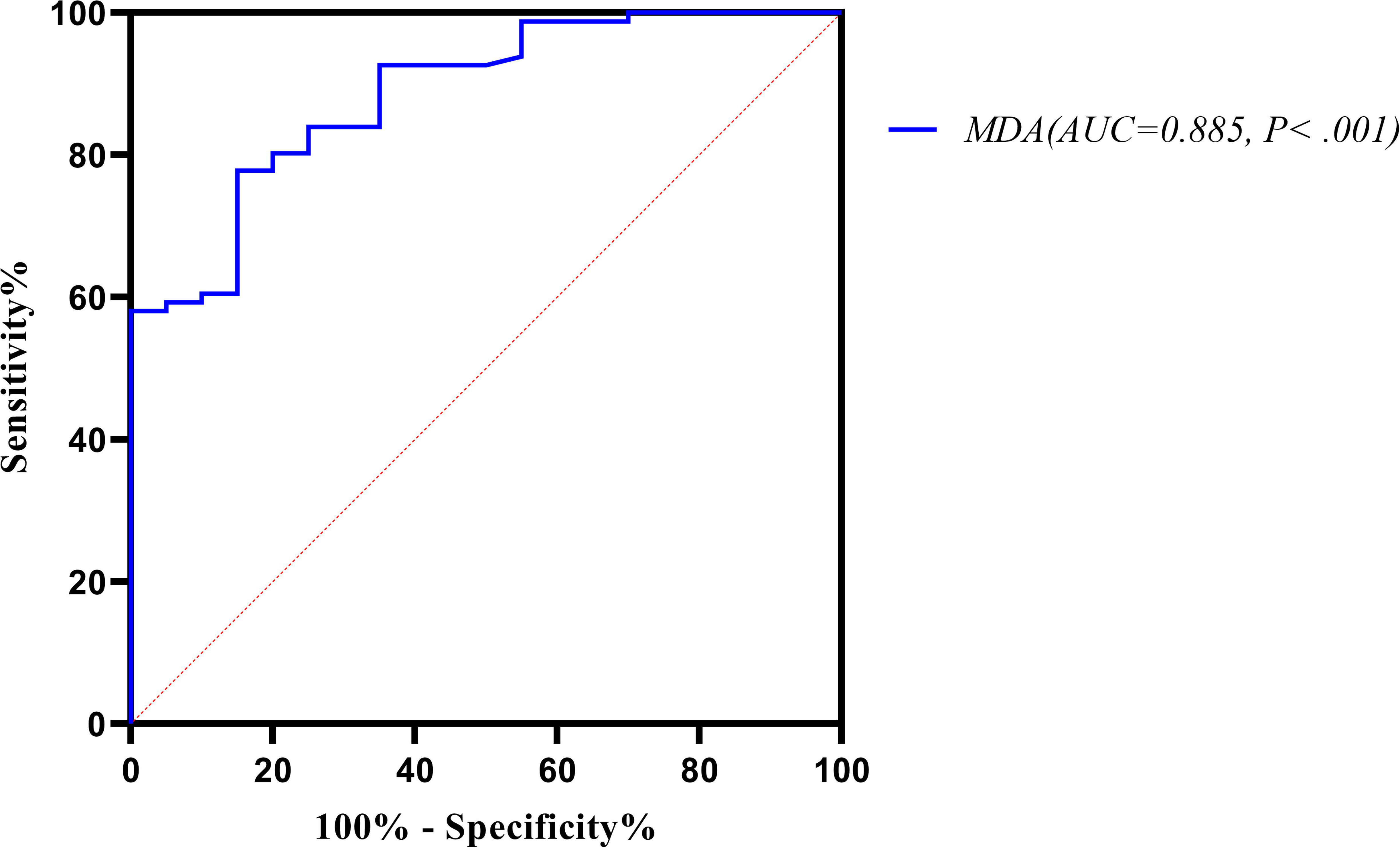
ROC curves of baseline MDA levels predicting treatment responses of SSRI-treated patients. ROC, receiver operating characteristic.

## 4. Discussion

This study explored SSRIs’ effects on peripheral oxidative stress markers in GAD patients. There were several significant findings from this research. First, MDA, cortisol, and LPO levels in GAD patients were higher than in HCs, while SOD, GSH-Px, and CAT levels were lower than in HCs. Second, the baseline levels of MDA, LPO, NO, and cortisol were positively correlated with anxiety severity, while GSH-Px was negatively correlated. Third, after eight weeks of SSRI treatment, GSH-Px levels tended to be upregulated, while MDA and LPO showed a downward trend. Furthermore, alterations in the MDA levels paralleled changes observed in the anxiety measurements. Finally, we determined that baseline MDA levels predicted treatment responses to SSRIs.

Our study revealed an oxidative stress imbalance in GAD patients. Elevated MDA, LPO, and cortisol levels indicated increased oxidative stress, and decreased SOD, GSH-PX, and CAT suggested weakened antioxidant mechanisms, which is consistent with the findings reported by Emhan et al.[30]. That study revealed that GAD patients exhibited a higher total oxidant status (TOS) and oxidative stress index (OSI), as well as a lower total antioxidant status (TAS), compared to healthy controls. Another study involving 14 individuals with post-traumatic stress disorder (PTSD) and 14 healthy participants revealed no alterations in the levels of MDA, SOD, CAT, and GSH-Px activity. This finding could be attributed to the body’s efficient capacity for rapid production of detoxifying free radicals[31]. The increase in oxidants and decrease in antioxidants observed in our study likely indicates that GAD patients experience oxidative stress, which impairs the antioxidant system, resulting in an insufficient antioxidant capacity. In addition, the oxidative stress marker levels in GAD patients at baseline were significantly correlated with GAD symptom severity. The oxidative stress products (MDA r=0.349, LPO r=0.470, and cortisol r=0.367) were positively correlated, and the antioxidant oxidase system (GSH-Px r=-0.458) was negatively correlated, which was consistent with previous studies[32, 33]. Increasing evidence indicates that the HPA axis, which mediates glucocorticoid secretion, is activated in stress responses[34]. Elevated glucocorticoid levels are associated with lipid peroxidation in neuronal cells, resulting in excessive accumulation of MDA, which can have toxic effects on neurotransmitters and inactivate membrane receptors, leading to a range of psychiatric disorders[35]. Mahmoodzadeh Y et al. [36] came to similar conclusions, reporting that MDA expression was higher in the hippocampus (HIP) and prefrontal cortex (PFC) during chronic, noise-induced depression and anxiety-like behavior in mice. The antioxidant N-acetylcysteine (NAC) can reduce MDA levels after administration. This antioxidant system is depleted in GAD patients, and H_2_O_2_ accumulation leads to mitochondrial DNA damage, further aggravating neuropathological changes in the brain[37].

The relationship between the antioxidant enzyme system and anxiety disorders is complex, and conflicting data have been reported. For example, Masood et al. [38]indicated that inhibition of glutathione synthesis could induce anxiety-like behavior by directly injecting L-butylthioneine -(S, R) sulfoxide into the mouse hippocampus. Thus, the Masood et al. study supports our results that GSH synthesis was reduced in GAD patients. Moreover, other studies have shown that SOD overexpression was positively correlated with anxiety symptoms in GAD patients; the increased SOD activity might be caused by the increased oxygen-free radical levels and the compensatory enhancement of enzymatic reaction against lipid peroxidation[39]. While elevated oxidative stress products consistently appear across all anxiety disorders, this is not the case for the antioxidant enzyme system. We believe that it is more likely that antioxidant mechanisms are diminished in GAD patients, as our results demonstrated. However, the interaction between these two factors needs additional study.

Several clinical studies have reported that SSRIs exhibit antioxidant properties in the treatment of neurological disorders such as anxiety, depression, and neurodegenerative diseases[40–42]. The results of this study revealed that after SSRI treatment, the MDA and LPO levels decreased in GAD patients, while GSH-Px levels increased. Thus, our results showed that the alleviation of anxiety symptoms might be linked to altered levels of oxidative stress markers. According to a meta-analysis conducted by Jimenez et al.[43], patients with depression exhibited increased serum MDA levels, decreased GSH-Px levels, and elevated red blood cell SOD levels, which were partially ameliorated by antidepressant treatment. Specifically, the MDA levels were reduced to that of healthy individuals, while the SOD level showed a declining trend without reaching normalcy, and no significant changes were observed in GSH-Px levels. The outcome discrepancies could be attributed to the pathological absence of up-regulation in the underlying antioxidant pathway.

It is essential to consider that our study focused on individuals with GAD, and variations in blood sample types should be considered. Some scholars argue that assessing the oxidative stress state of red blood cells may offer a more accurate reflection of cerebral conditions[44], thus providing us with a valuable research strategy. Indeed, animal experiments have indirectly verified our findings. Shahzad et al.[45] found that atenolol combined with escitalopram pre-treatment reduced MDA levels in rat brain tissue. Furthermore, anxiety-related behavior in the elevated plus maze (EPM) revealed that such treatment significantly increased the preference percentage, the number of open arm entries, and the open arm time. It has also been reported that the SSRI fluoxetine increased antioxidant capacity, which was measured by increasing the activity of CAT and GSH-Px in the hippocampus of rats[46].

Overall, our research has yielded promising insights that the alleviation of anxiety symptoms following treatment with SSRIs may be associated with alterations in levels of oxidative stress markers. However, the mechanism has yet to be identified. Lukic et al. [47] reported that SSRIs activated Nrf_2_ expression and upregulated the transcription of various antioxidant target genes and mitochondrial biogenesis-related genes. However, further research is needed to determine whether the Nrf2 signaling pathway activation is required for SSRIs to exert antioxidant effects.

To our knowledge, this is the first study to demonstrate how oxidative changes correlate with changes in anxiety after SSRI treatment in patients with GAD, indicating that patients with a greater reduction in MDA levels exhibit greater improvement in anxiety. Because MDA was associated with anxiety symptoms observed at the baseline and with treatment responsiveness to SSRIs, it is logical to continue investigating the predictive value of baseline MDA for treatment response to SSRIs. These studies should have substantial clinical significance.

The prediction of treatment outcomes using biomarkers also will benefit personalized medicine. Our study revealed that patients with lower baseline MDA levels exhibited better SSRI treatment responses. These observations parallel the study conducted by Buosi et al. [48], who found that poor treatment responses in schizophrenia patients were associated with high lipid peroxidation levels. The ROC curve also demonstrated that baseline MDA levels served as a reliable predictor of SSRI responses in GAD. Increases in lipid peroxides in the brain are associated with numerous neuropsychiatric diseases. MDA is the end-product of lipid peroxidation, and its toxicity directly reflects the degree of oxidative stress damage in neurons[49–51]. Consequently, it is reasonable to conclude that lower baseline MDA levels might predict a better treatment outcome in GAD patients treated with SSRIs. The mechanism of action of SSRIs is to elevate the synaptic concentrations of 5-HT, thereby exerting an antidepressant effect. The augmentation of 5-HT neurotransmitter activity can suppress microglial activation, expedite ROS elimination, and diminish MDA generated by membrane lipid peroxidation. It also diminishes the body’s utilization of antioxidants[52]. Studies have documented that prolonged treatment with 10 mg/kg fluoxetine can augment hippocampal GSH levels in mice[53].

However, additional investigations are necessary to validate these findings and provide a more individualized approach to treating GAD patients.

There are several limitations associated with this study. First, 101 patients finally completed this study, and 54 patients exhibited remission after eight weeks of treatment. Because a smaller sample size would adversely affect statistical power, we did not conduct a dichotomous distinction of “remitter” vs. “non-remitter.” Notably, “responders” and “non-responders” meet the widely accepted definition of clinically meaningful but not necessarily complete improvement. Therefore, the dichotomy between “remitters” and “non-remitters” is more comprehensive in distinguishing treatment outcomes, and patients are less likely to relapse. Thus, future studies should be conducted utilizing a larger sample size. Second, although we attempted to control for the influence of depression, the presence of depression could not be ruled out in our study due to the high prevalence of comorbid generalized anxiety disorder and depression. Kalin et al.[54] reported that lifetime comorbidity for GAD and depression is 43%. Furthermore, the diagnostic criteria for GAD indicate a class associated with major depressive disorder[55]. Therefore, more stringent inclusion criteria might be helpful in future studies. Third, numerous studies suggested that the development of mental illnesses can trigger oxidative stress and inflammation via oxidation and nitrosation pathways, leading to DNA damage and the expression of DNA damage markers (8-OHdG)[56–58]. These markers might be more reliable indicators of oxidative stress than enzymes and should be considered in future research. Finally, we measured biomarkers peripherally and not in the CNS. Whether oxidative stress markers are regulated in the CNS in the same way as in the periphery has been questioned[59] and needs further clarification.

## 5. Conclusion

This study provided new evidence concerning how oxidative stress is involved in the pathophysiology of GAD. This study also demonstrated that decreased baseline MDA levels might predict the treatment response to SSRIs. Thus, the therapeutic potential of targeting the oxidative stress system in psychiatry is noteworthy. Although further research on GAD is warranted, current studies have revealed promising outcomes.

## Acknowledgement

We want to thank all our participants who made this study possible.

## 6. Statement of Ethics

This study was approved by the Ethics Committee of Huzhou Third Municipal Hospital (approval no. 2021-017). All procedures used in this study involving human participants were carried out in accordance with the World Medical Association Declaration of Helsinki (revised in 2013). After receiving a complete description of the study protocol, written informed consent was obtained from all participants prior to beginning the study.

## 9. Author contributions

Lijun Cui and Jingjing Lu contributed to the writing of the manuscript. Zhongxia Shen, Shenliang Yang, and Shikai Wang contributed to the medical treatment of the patients. Jielin Zhu contributed to data collection and assembly. Huanxin Chen and Xinhua Shen completed the study design. All authors have read and agreed to the published version of the manuscript.

## 10. Data Availability Statement

Data are not publicly available due to ethical considerations. Further inquiries can be directed to the corresponding author.

